# A protracted cholera outbreak in Nairobi City County associated with mass gathering events, Kenya, 2017

**DOI:** 10.1101/2023.11.20.23298754

**Authors:** Philip Ngere, Daniel Langat, Isaac Ngere, Jeanette Dawa, Emmanuel Okunga, Carolyne Nasimiyu, Catherine Kiama, Peter Lokamar, Carol Ngunu, Lyndah Makayotto, M. Kariuki Njenga, Eric Osoro

**Author notes:** Corresponding author (PN).

## Abstract

Cholera continues to cause many outbreaks in low and middle-income countries due to inadequate water, sanitation, and hygiene services. We describe a protracted cholera outbreak in Nairobi City County (NCC), Kenya in 2017. We reviewed the cholera outbreak line lists from NCC in 2017 to determine its extent and factors associated with death. A suspected case of cholera was any person aged >2 years old who had acute watery diarrhea, nausea, or vomiting, whereas a confirmed case was where *Vibrio cholerae* was isolated from the stool specimen. We summarized cases using for continuous variables and proportions for categorical variables. Associations between admission status, sex, age, residence, time to care seeking, and outbreak settings; and cholera associated deaths were assessed using odds ratio (OR) with 95% confidence interval (CI). Of the 2,737 cholera cases reported, we analyzed 2,347 (85.7%) cases including 1,364 (58.1%) outpatients, 1,724 (73.5%) not associated with mass gathering events (MGEs), 1,356 (57.8%) male and 2,202 (93.8%) aged ≥5 years, and 35 deaths (case fatality rate: 1.5%). Cases were reported from all the Sub Counties of NCC with an overall county attack rate of 50 per 100,000 people. *Vibrio cholerae* Ogawa serotype was isolated from 78 (34.8%) of the 224 specimens tested and all isolates were sensitive to tetracycline and levofloxacin but resistant to amikacin. The odds of cholera-related deaths was lower among outpatient cases (aOR: 0.35; [95% CI: 0.17-0.72]), age ≥5 years old (aOR: 0.21 [95% CI: 0.09-0.55]), and MGEs (aOR: 0.26 [95% CI: 0.07-0.91]) while threefold higher odds among male (aOR: 3.04 [95% CI: 1.30-7.13]). NCC experienced a protracted and widespread cholera outbreak with a high case fatality rate in 2017.

**Author Summary:** Cholera outbreaks are common in Kenya. The highest number of cases were reported in 2015 with 10,536 cases across the country. The subsequent three years also recorded a high number of cholera cases with 6,137 cases in 2016, 4,217 cases in 2017, and 5,638 cases in 2018. During the four years (2015-2018), NCC contributed less than 20.0% of the cholera cases, except in 2017 when the county reported 56.9% (2,737) of the cases. In this study, we sought to describe the burden of cholera in NCC during the 2017 outbreak. We reviewed the cholera outbreak data from NCC in 2017 to determine its extent and risk factors for cholera-related deaths. The findings depict a prolonged and widespread cholera outbreak with the likelihood of death higher among male cases but lower among outpatients, cases aged ≥5 years old, and cases from the MGEs. More studies on the factors associated with cholera-related deaths are necessary to inform public health response.

## Introduction

Cholera, an acute watery diarrheal (AWD) disease, poses a significant threat to public health worldwide. With a potential fatality rate of up to 5% in untreated cases, cholera remains a public health concern, requiring comprehensive research and evidence-based interventions [1]. The causative agent, *Vibrio cholerae*, is transmitted through the feco-oral route, either directly from person-to-person or through contaminated fluids, food, flies, and fomites [2]. Cholera outbreaks are intrinsically linked to poor water, sanitation, and hygiene (WASH) indicators, affecting both households and communities [4]. Notably, densely populated areas such as informal settlements, displaced persons’ camps, and mass gathering events (MGEs) serve as hotspots for cholera transmission due to heightened person-to-person interactions and increased strain on WASH facilities [5,6].

Globally, cholera continues to exact a heavy toll, with an estimated 1.3 billion people at risk and millions of cases and tens of thousands of deaths reported each year [7]. Sub-Saharan Africa shoulders the greatest burden of cholera, accounting for up to 83% of deaths reported between 2000 and 2015 [8]. Kenya, situated in this high-risk region, has experienced a fluctuating cholera epidemiology, with endemicity influenced by varying WASH indices. The country’s commitment to ending cholera by 2030, as supported at the 71^st^ World Health Assembly in 2018, highlights the urgency in addressing this persistent health challenge.

Kenya’s experience with cholera dates to its first detection in 1971, with large outbreaks in 1997-1999 and 2007-2010. Subsequently, cases declined in 2011-2014, only to re-emerge in 2015 [9,10]. Presently, cholera in Kenya is endemic and is characterized by regional disparities in disease incidence [14]. Nairobi City County (NCC), the country’s capital, has played a significant role in this epidemiological landscape. In 2015, Kenya recorded its highest annual cholera case count, with 10,536 cases reported nationwide, including 1,845 (17.5%) cases within NCC. In the subsequent three years (2016-2018), Kenya consistently reported high cholera caseloads, exceeding 4,000 cases annually, with 2017 marking a surge in NCC, contributing to 2,737 (64.9%) of the reported 4,217 cases countywide. Here we characterize the 2017 cholera outbreak in NCC and identify risk factors associated with mortality.

## Methods

### Study design

We conducted a cross-sectional study of the cholera outbreak from Nairobi City County, Kenya, between 1^st^ January to 31^st^ December 2017.

### Study area

The study was conducted in Nairobi City County (NCC), the capital city of and one of 47 counties in Kenya. As of 2017, NCC had a population of 4,697,274 with a population density of 6,949 persons per square kilometer [13]. NCC comprises 17 administrative units (sub-counties) and is characterized by diverse settings, including formal settlements and informal settlements. Over 60% of NCC’s population resides in informal settlements, which occupy only 6% of the land area [16].

### Cholera outbreak detection

Cholera cases were defined based on Kenya’s Integrated Diseases Surveillance and Response (IDSR) technical guidelines. A suspected case of cholera was defined as any person aged two years or older who presented with AWD during the outbreak period. A confirmed case was any suspected case from whose stool or rectal swab specimens *Vibrio cholerae* was isolated. Any suspected case that was epidemiologically linked to a confirmed case or from whose stool or rectal swab specimens test by a rapid diagnostic test turned positive for *Vibrio cholerae* was considered a probable case [16]. Upon confirmation of a cholera outbreak (one confirmed case), probable cases were treated as confirmed cases but underwent weekly random sampling and testing to confirm continuity of the outbreak and monitor the *Vibrio cholerae* strains [17].

### Data source

The data for this study were sourced from cholera outbreak line lists for NCC, collected by the NCC public health surveillance officers during the outbreak period. These line lists are comprehensive records of cholera cases, including variables such as age, sex, admission status, outbreak settings (including MGEs), dates of hospital visits, dates of symptom onset, disease outcomes, pathogen strains, and antimicrobial susceptibility test results. A mass gathering event (MGE) was a function attended by many people that could strain the health systems beyond the capacity provided e.g., sporting, religious, and cultural events activities. The confirmatory stool or rectal swab specimens collected from suspected cases to confirm the cause of the AWD were transported in Cary-Blair transport media to the National Public Health Laboratories (NPHL) and plated on thiosulfate-citrate-bile salts agar (TCBS). The resultant colonies were evaluated using biochemical reactions and serotyped using commercial antisera. Colonies were also tested for antimicrobial susceptibility using disk diffusion method.

### Data analysis

All cases listed in the cholera outbreak line lists for NCC between January 1, and December 31, 2017, were included. To prepare the data for analysis, a thorough data cleaning process was undertaken to address inconsistent data formats, repeated entries, and missing or inconsistent data points using Microsoft Excel. Data was analyzed in Epi Info statistical software. Summary tables with means for continuous variables and proportions for categorical variables were generated. Attack rate (AR) was the number of reported cholera cases per 100,000 population. Case fatality rate (CFR) was the percentage of deaths among all cases. The antimicrobial susceptibility tests results were summarized as proportions and presented using stacked bar charts.

In the bivariate analysis, the association between several variables (admission status, sex, age, residence, time to care seeking, and outbreak setting) and cholera-related death was assessed and crude odds ratios (cOR) with 95% confidence intervals (CI) presented. Variables with p≤0.20 were included in an unconditional multivariable logistic regression model using the backward elimination method and the adjusted ORs reported. Statistical significance was set at p <0.05.

### Ethical considerations

Approval to access the line list data was sought from the Kenya Ministry of Health’s Division of Disease Surveillance and Response. The study used de-identified data that had been collected during outbreak response activities and therefore informed consent was not sought.

## Results

From an initial line list of 2,737 from Nairobi City County, we analyzed 2,347 cases after excluding 339 (14.1%) due to misclassified dates, 36 (1.5%) duplicates, and 15 (0.6%) incomplete records. The median age of cases was 31.0 years, ranging from 0.5 to 86 years. Many cases (93.8%) were aged ≥5 years, with 80.4% falling within the working age group of 21-65 years. Of the cases, 57.8% were male, and 58.1% were outpatients while 26.5% of cases were linked to MGEs. The overall case fatality rate (CFR) was 1.5%, with higher rates observed in children <5 years old (4.8%), cases from other counties (2.6%), and inpatients (2.3%) (**Table 1**)

**Table 1:** Distribution of mortalities from cholera in Nairobi City County, Kenya in 2017.

|  | All cases |  | Deaths |  |  |
| --- | --- | --- | --- | --- | --- |
| Variable | n | % | n | % | CFR (%) |
| Admission status |  |  |  |  |  |
| Outpatient | 1,364 | 58.1 | 12 | 34.3 | 0.9 |
| Inpatient | 983 | 41.9 | 23 | 65.7 | 2.3 |
| Sex |  |  |  |  |  |
| Male | 1,356 | 57.8 | 28 | 80.0 | 2.1 |
| Female | 991 | 42.2 | 7 | 20.0 | 0.7 |
| Age (years) |  |  |  |  |  |
| <5 | 145 | 6.2 | 7 | 20.0 | 2.1 |
| ≥5 | 2202 | 93.8 | 28 | 80.0 | 0.9 |
| Residence |  |  |  |  |  |
| Other Counties | 231 | 9.8 | 6 | 17.1 | 2.6 |
| Nairobi County | 2,116 | 90.2 | 29 | 82.9 | 1.4 |
| Care seeking (hours) |  |  |  |  |  |
| ≥ 24 | 1,576 | 67.1 | 24 | 68.6 | 1.5 |
| < 24 | 771 | 32.9 | 11 | 31.4 | 1.4 |
| Outbreak setting |  |  |  |  |  |
| Mass gatherings events | 623 | 26.5 | 3 | 8.6 | 0.5 |
| Non-mass gathering events | 1,724 | 73.5 | 32 | 91.4 | 1.9 |
| Total (NCC) | 2,347 | 100.0 | 35 | 100.0 | 1.5 |

Cholera cases were reported during 41 (78.8%) of 52 epidemiological weeks. Continuous reporting of cases was observed from epidemiological week 14 through to week 48. An increase in the number of cases was evident between weeks 28 and 45. Two distinct peaks were identified during weeks 29-30 and weeks 35-36, which were attributed to events with MGEs. A MGE was associated with a large increase in cases during week 25 (**Figure 1**).

**Figure 1:**
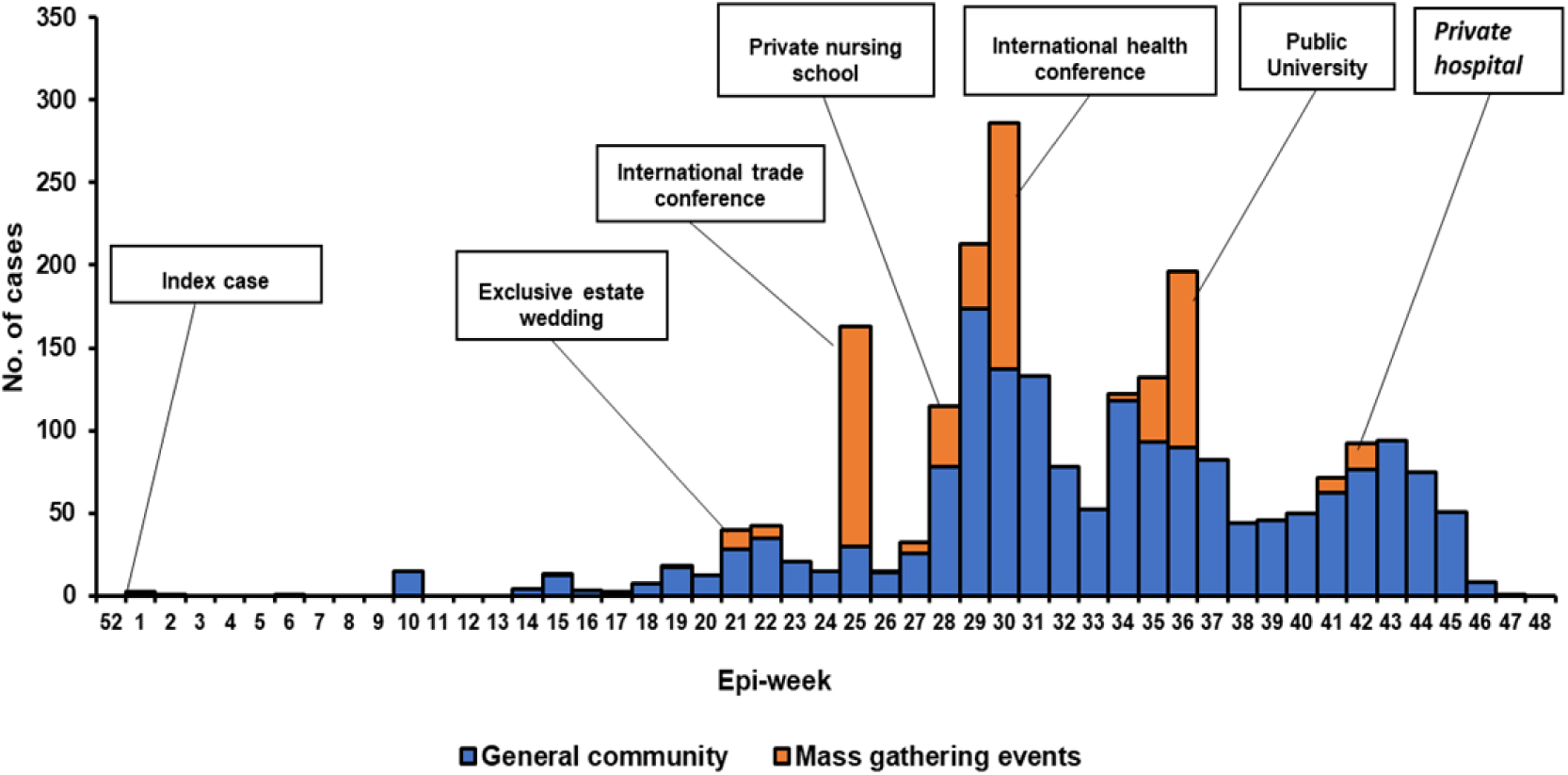
Epi-curve of cholera cases reported by week in Nairobi City County, Kenya, 2017.

All the sub counties in NCC reported cholera cases with an overall county AR (cases per 100,000 population) of 50.0. Three sub counties recorded ARs of ≥100: Embakasi East (243.4), Embakasi West (102.7), and Langata (100.0) while those with the highest CFRs were Mathare (7.1%), Embakasi North (5.9%), and Kasarani (4.0%) (**Figure 2**).

**Figure 2:**
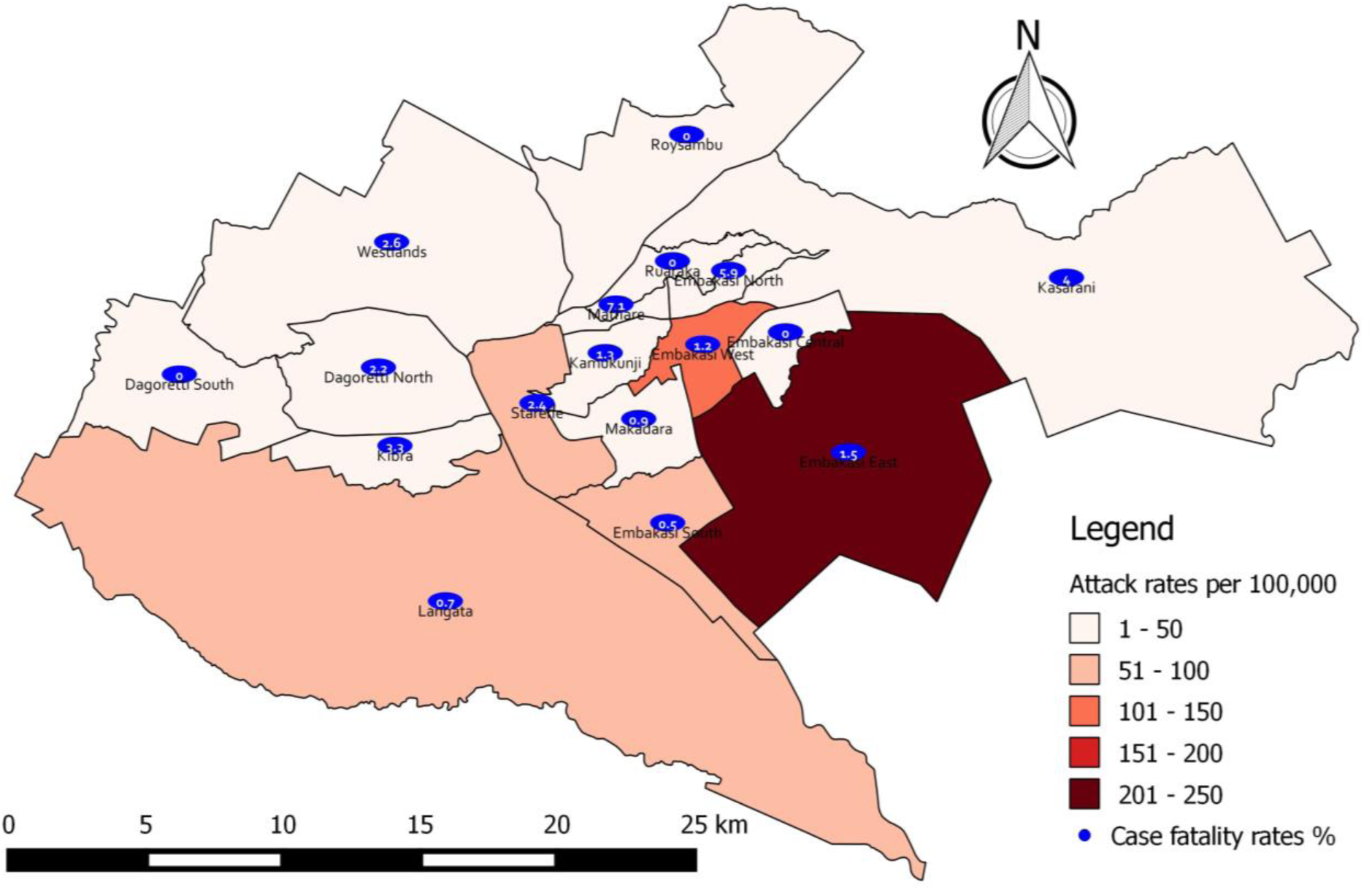
Cholera attack rate per 100,000 population and case fatality rates (%) by Sub Counties in Nairobi City County, 2017.

Of the 224 rectal swab or stool samples that underwent culture and serotyping, *Vibrio cholerae* of the Ogawa serotype was identified in 78 (34.8%). All the isolates were sensitive to tetracycline and levofloxacin but resistant to amikacin (**Figure 3**).

**Figure 3:**
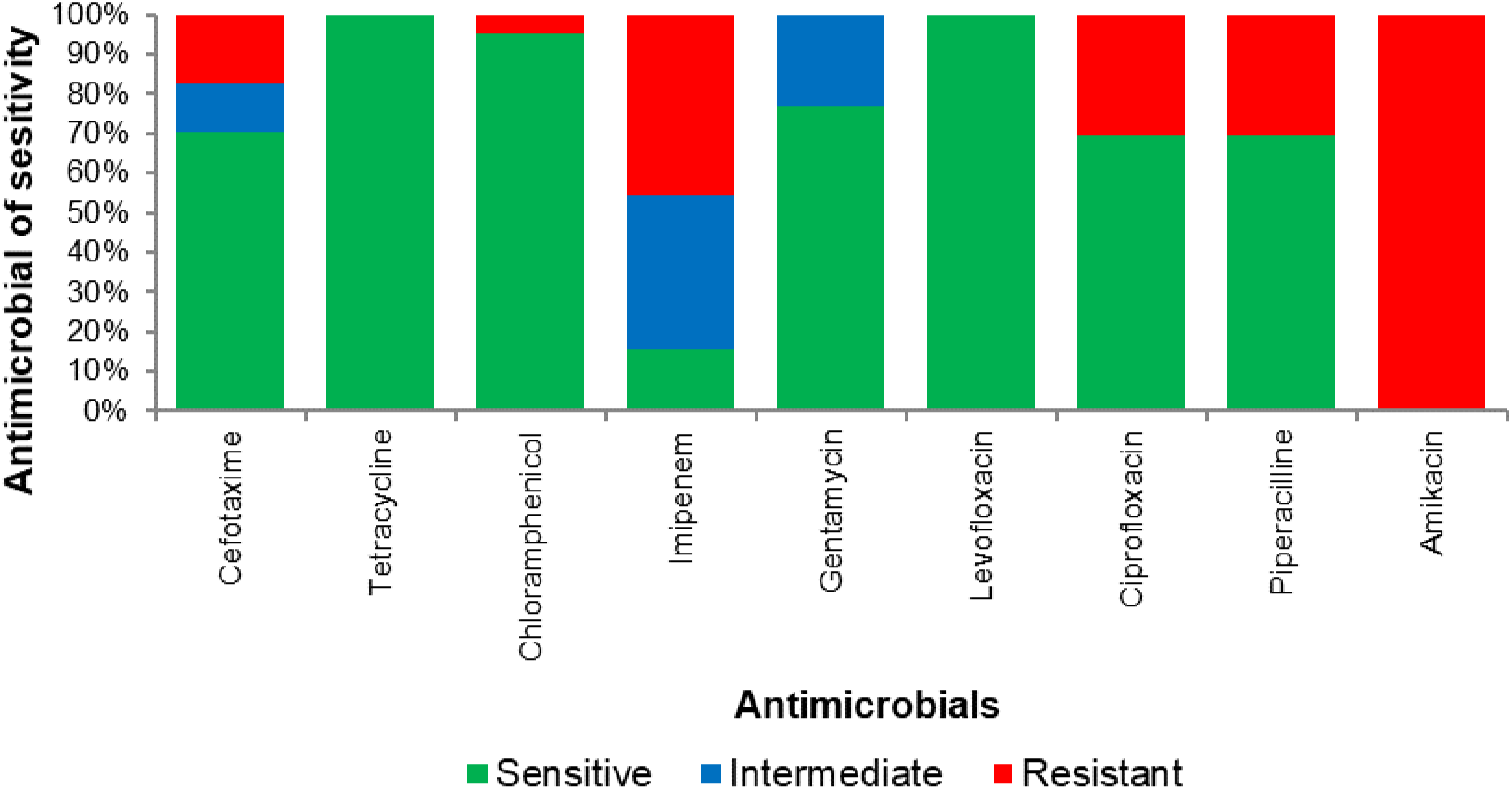
Levels of Vibrio cholerae antimicrobial sensitivity by type of antimicrobial from Nairobi City County, 2017.

On bivariate analysis, the odds of a cholera-related death was higher among male (cOR: 2.83 [95% CI: 1.21-6.62]). There was 63% lower odds of cholera-related deaths among outpatients compared to inpatients (cOR: 0.37 [95% CI: 0.18-0.76]). Other factors associated with lower odds included being aged <5 years old (cOR: 0.24 [95% CI: 0.10-0.58]), seeking medical attention within the 24 hours of symptom onset (cOR: 0.61 [95% CI: 0.28-1.33]), and cases linked to MGEs (cOR: 0.20 [95% CI: 0.05-0.75] (**Table 2**). In the multivariable logistic regression analysis, factors that were independently associated with lower odds of cholera associated cholera-related deaths included outpatient cases (adjusted Odds Ratio [aOR]: 0.35; 95% CI: 0.17-0.72), age ≥5 years olds (aOR: 0.21 [95% CI: 0.09-0.55]), and linkage to MGEs (aOR: 0.26 [95% CI: 0.07-0.91]). Conversely, male had threefold higher odds of cholera-related death (aOR: 3.04 [95% CI: 1.30-7.13]) (**Table 2**).

**Table 2:** Factors associated with cholera deaths in Nairobi City County, Kenya in 2017.

| **Variable** | **Bivariable analysis** |  | **Multivariable analysis** |  |
| --- | --- | --- | --- | --- |
|  | **cOR (95% CI)** | **p-Value** | **aOR (95% CI)** | **p-Value** |
| **Admission status** |  |  |  |  |
| Outpatient | 0.37 (0.18-0.76) | 0.007 | 0.35 (0.17-0.72) | 0.004 |
| Inpatient | Ref |  | Ref |  |
| **Sex** |  |  |  |  |
| Male | 2.83 (1.21-6.62) | 0.017 | 3.04 (1.30-7.13) | 0.010 |
| Female | Ref |  | Ref |  |
| **Age (years)** |  |  |  |  |
| ≥ 5 | 0.24 (0.10-0.58) | 0.001 | 0.21 (0.09-0.55) | 0.001 |
| < 5 | Ref |  | Ref |  |
| **Residence** |  |  |  |  |
| Other Counties | 2.16 (0.81-5.73) | 0.123 | 2.35 (0.90-6.13) | 0.080 |
| Nairobi County | Ref |  | Ref |  |
| **Care seeking (hours)** |  |  |  |  |
| ≥ 24 | 0.61 (0.28-1.33) | 0.213 | 0.56 (0.26-1.21) | 0.142 |
| < 24 | Ref |  | Ref |  |
| **Outbreak setting** |  |  |  |  |
| Mass gatherings events | 0.20 (0.05-0.75) | 0.017 | 0.26 (0.07-0.91) | 0.035 |
| Non-mass gatherings events | Ref |  | Ref |  |

cOR - crude Odds Ratio; aOR - adjusted Odds Ratio

## Discussion

In 2017, Nairobi City County (NCC) experienced a large cholera outbreak which was protracted and widespread with a propagated transmission pattern accentuated by point source outbreaks during mass gathering events (MGEs). This substantially contributed to Kenya’s cholera cases which can be attributed to NCC’s vast informal settlements which are characterized by poor WASH indicators and overcrowding that are known drivers of cholera outbreaks [18]. The influx of refugees escaping from designated camps in northern Kenya due to security threats; inadequate education and medical services; limited livelihood opportunities; and harsh climatic conditions together with the increase in the day-population due to migrant workers may have further exacerbated the situation [19,20]. These dynamics, coupled with spillovers of cases from neighboring counties with cholera outbreaks in search of care, likely sustained the protracted nature of this outbreak.

The outbreak had an overall case fatality rate (CFR) higher than the <1.0% stipulated by WHO with timely and appropriate management [21]. Specific demographics and settings exhibited even higher CFRs. The identified risk factors for cholera-related mortalities included being male, inpatient status, age <5 years old, and non-MGE linked settings. Whereas the high CFR among inpatient cases observed in this study is indicative of potential gaps in case management, demonstrated delays in care seeking may have contributed to worsening the disease outcomes [22,23]. The gender and age variations in cholera cases observed in this study can be attributed to societal gender roles. Majority of cases were male contrary to previous studies [24,25]. While cholera infection has no sex predilection, female sex has been shown in other studies to be disproportionately more affected during cholera outbreaks. The higher numbers of male cases reported in this cholera outbreaks may be attributed to transmission occurring mostly out of the households. This is further reaffirmed by the high transmission within the age group 21-65 years as opposed to the young (ages <5 years) and elderly (ages >65 years) suggestive of work settings transmission [26,27]. The high CFRs among male cases is consistent with other studies that have demonstrated female sex as protective against unfavorable cholera outcomes [26,27]. This has been attributed to men seeking healthcare late and less often [28]. This study also concurred with previous observations of high cholera CFRs among children [29,30].

The sub counties that are hosts to major informal settlements in NCC such as Mathare, Embakasi North, Kasarani, Kibra, Starehe, and Dagoreti North registered highest CFRs. The disparities in CFRs across sub counties and settings underscore the influence of socio-economic factors, WASH infrastructure, and health services accessibility on cholera outcomes [31]. Typically, few cholera outbreaks would be expected within formal settlements with good sanitation and safe water [32]. Interestingly, this study observed explosive point source outbreaks linked to MGEs within formal settlements. This paradox can be explained by the introduction of infection to the institutions or MGEs through the practice of outsourcing ready-to-eat foods and drinks that are likely contaminated or contamination of institutional foods by infected food handlers, cross contamination from infected residents or guests [33]. Hypothetically, access to good sanitation and safe water in these settings implies that the population has minimal or no exposures to *Vibrio cholerae* hence low level or no immunity against cholera. In the absence of any routine cholera vaccination, a spill over into this population would then lead to explosive point source outbreaks [34,35]. These explosive point source outbreaks linked to the MGEs, however, did not register commensurate CFRs possibly due to better WASH and access to health services within the formal settlements.

Cholera outbreaks have been shown to coincide with onset of rainfall [36]. The cholera outbreaks in Nairobi City County in 2017 presented major peaks in epi-weeks 25, 30, and 36; a deviation from the rainfall peaks expected from March to May (epi-weeks 9-22) and October to December (epi-weeks 40-52) [37]. The peaking pattern of cholera cases as noted in this study could be attributed to explosive point source outbreaks during the MGEs in formal settlements between epi-weeks 22 and 36.

The isolation of *Vibrio cholerae* of the Ogawa serotype and its antimicrobial susceptibility patterns align with previous observations and have implications for treatment strategies [38]. However, the emerging resistance to second-line antimicrobials presents a potential challenge for _cholera control_ [39]. Whereas this study shows complete susceptibility to tetracycline which is a first line drug used in treatment and newer fluoroquinolones, widespread resistance is exhibited to ciprofloxacin, a commonly used second line drug alongside third generation cephalosporins, *b*-Lactam penicillins and aminoglycosides. This presents a possible emerging challenge to the control of cholera outbreak as witnessed in other settings [40].

## Limitations

A limitation of study was the reliance on reported outbreak data, which might not capture the actual cholera burden. Additionally, the calculated attack rates, based on census reports, might underestimate the actual population at risk, given the city’s dynamic day-population.

## Conclusions

The 2017 cholera outbreak in NCC was marked by protracted and widespread transmission, predominantly within informal settlements, with sporadic explosive outbreaks during MGEs. The outbreak underscores the need for a multifaceted approach to cholera control. Enhanced surveillance, especially during mass gathering events, is paramount. Addressing the challenges unique to informal settlements, such as improving water, sanitation, and hygiene facilities, is crucial. Public health campaigns should prioritize raising cholera awareness in high-risk areas to promote early detection and treatment. Given the potential for spillovers from neighboring counties, a coordinated inter-county response is essential. Lastly, further research into the specific factors driving cholera transmission during such outbreaks can provide actionable insights for future prevention measures.

## Authors’ contributions

PN, DL, IN, JD, EO, CN, CK, PL, CN, cleaned, merged, and coded the line lists under the guidance of LM, MKN, and EO. All authors participated in data analysis and report writing. PN drafted the manuscript with input from all the authors. All the authors have reviewed and approved the final version of the manuscript.

## Ethics statement

Ethical review and approval were waived for this analysis based on retrospective data.

## Data availability statement

All the data underlying the findings described are available within the manuscript.

## Acknowledgements

We acknowledge the Ministry of Health, Kenya, and the Nairobi City County Government for providing access to the cholera outbreak line lists used in this study, authorizing the study, and allowing its publication.

## Disclaimer

The conclusions reached in this paper are for the authors and do not necessarily represent the position of the Ministry of Health, Kenya.

